# Signs and symptoms associated with a need for supervision in patients with Alzheimer’s disease

**DOI:** 10.1101/2022.10.20.22281336

**Authors:** Celia C. Huey, Anton Kociolek, Kayri K. Fernandez, Michelle Hernandez, Reena T. Gottesman, Megan Barker, Stephanie Cosentino, Yaakov Stern

## Abstract

Caregivers identify the need for continuous supervision of Alzheimer’s disease patients while awake as a “milestone” functional change that has a profound impact on the lives of the caregiver and patient. The specific predictors of this important functional change are not known. We determined specific cognitive, neuropsychiatric, and motor symptoms associated with Alzheimer’s disease patients needing supervision while awake in a longitudinal, ethnically diverse cohort of Alzheimer’s disease patients in Northern Manhattan. At the initial visit, neuropsychiatric and behavioral symptoms, including more hallucinations (OR=3.12) and lower elation (OR=0.13) were associated with the need for supervision while awake, as were poor memory (OR=0.89) and higher letter fluency (OR=1.33) abilities. The findings presented in the current study can aid clinicians and caregivers in prognosis and planning, suggest symptoms to target with non-pharmacological and pharmacological treatments to forestall this important functional “milestone”, and suggest priorities for future research.

**Significance Statement:**

- Need for supervision while awake is identified by caregivers of AD patients as a “milestone” symptom that greatly changes caregiving needs and quality of life
- Specific neuropsychiatric and cognitive symptoms, including hallucinations, elation, and memory and executive dysfunction, are associated with a significantly increased or decreased need for supervision while awake
- These patient-centered findings aid prognosis and planning, and indicate symptoms to prioritize for treatment and future research to improve AD patient functioning

## Introduction

Over 6 million people in the United States suffer from dementia due to Alzheimer’s disease (AD) and more than 16 million family and other unpaid caregivers provide an estimated 18.6 billion hours of care annually to patients with AD and related dementias (ADRD) (1). The majority of care for patients with ADRD is provided at home (80% of patients) by unpaid caregivers (83% of caregivers) (2). AD is characterized by functional impairment, however, there is only a moderate correlation between functional and cognitive changes (3). Caregivers report that the functional changes of AD are often more difficult to manage than the cognitive symptoms (4).

Previous research has established various risk factors that are associated with more severe disease outcomes, such as faster progression, higher financial burden, and out-of-home placement. The presence of extrapyramidal signs is associated with more rapid progression of Alzheimer’s disease and increased mortality (5, 6). Functional impairment and neuropsychiatric symptoms of AD are associated with increased financial burden. A one point increase on the Blessed Dementia Rating Scale, a functional measure, was associated with an increased cost of $1411 (7, 8), and depressive symptoms in AD were associated with an increased cost in medical expenses (9). Out-of-home placement is associated with a large increase in the cost of care (10). Several studies have examined the symptoms and functional impairments associated with out-of-home placement in persons with dementia, possibly in part because this is a clear change of patient status associated with increased cost of care that can be determined from records [see (11, 12) for systemic reviews and meta-analyses of these studies]. Summarizing over 25 of these studies, the authors of one review found that nursing home admission was most strongly associated with poorer cognition and to a lesser extent with behavioral and neuropsychiatric symptoms (11). Nursing home admissions were also associated with acute medical events, including hip fracture (11). Nursing home placement was also found to be associated with patient race and ethnicity with greater placement of White compared to African-American and non-white Hispanic patients, cognitive impairment, and the presence of one or more difficult behaviors (12). However, placement in an out-of-home setting is influenced by many factors in addition to the symptoms and functional impairment of the patient. These include the family financial situation, the availability of paid and unpaid home caregivers, need for insurance that supports out-of-home placement (e.g. Medicaid), availability of local out-of-home placement options, differences between countries, and cultural and individual attitudes about out-of-home placement, to name a few (13).

Need for continuous daytime supervision has been identified in interviews with caregivers as a “milestone” clinical symptom that greatly increases caregiver effort and burden (14, 15). The predictors of this important functional outcome have been minimally examined, possibly because this outcome must be ascertained from caregiver interviews rather than from records. The Predictors 3 study longitudinally follows an ethnically diverse community sample of patients and their caregivers in New York City (16). Participants receive extensive neuropsychological, neurological, and neuropsychiatric characterization. Among other measures, caregivers are administered the Dependence Scale (DS), a measure of functional and caregiving outcomes completed by a trained rater based on a caregiver interview (13, 17). The majority of other scales of dependence measure outcomes of increased care, such as out-of-home placement (13). The DS has shown a strong correlation with other measures of disability such as the Disability Assessment for Dementia (13, 18, 19). Previous studies have found that the baseline level of dependence on the DS was significantly, but weakly, correlated with the MMSE (20), CDR (21), memory and visuospatial ability (22), and factors of the Blessed Dementia Rating Scale (8, 17). Other cross-sectional studies have shown a significant correlation between total DS score and the MMSE, Clinical Dementia Rating Scale Sum of Boxes (CDR-SB) (21), and a weaker association with the Neuropsychiatric Inventory (23) and the Columbia University Scale of Psychopathology in Alzheimer’s disease (CUS-PAD) (24, 25). In the present study we examined the neuropsychological, neurological, and neuropsychiatric predictors of the need for the AD patient to be supervised while awake in a multi-ethnic, community-based, longitudinal sample, as reported by the primary caregiver. The goals of the study were to provide clinicians and caregivers with information on signs symptoms associated with this important functional outcome to: 1. Aid in prognosis and planning; 2. Target these symptoms for treatment in clinical settings; 3. Prioritize the development of improved treatments for these symptoms; 4. Provide directions for future research on this topic.

## Methods

### Participants

The current sample was drawn from the Predictors 3 cohort’s baseline visit. This is a community-based study of ethnically diverse participants with mild AD (CDR=1) who are assessed every 18 months with neuropsychological testing, and clinical and functional evaluations. The design and methods of this study have been described (16). Recruitment of the Predictors 3 cohort started in 2011 and is ongoing. For this study, we utilized data from Predictors 3 participants with an AD diagnosis (26) and who had an informant who could provide information on the functional status of the participant. Data from the first Predictors 3 visit at study entry was used in the analysis. Written informed consent was obtained and all study procedures were reviewed and approved by the New York State Psychiatric Institute Institutional Review Board.

### Measures and Procedures

The full list of Predictors 3 measures has been previously described (16). Due to the large numbers of neuropsychological tests administered in Predictors 3, a research neuropsychologist (M.B.) selected representative tests of major cognitive domains from the larger battery to analyze in the current study: Selective Reminding Test (SRT) Total Recall (Immediate) (27), Boston Naming Test Total (28), CFL letter fluency mean, and Benton Matching score (29). To evaluate extrapyramidal signs (EPS), selected items from the Unified Parkinson’s Disease Rating Scale were used comprising ratings of voice, facial immobility, resting tremor, rigidity (neck and each limb), brady/hypokinesia, posture, and gait on a 0 to 4 scale (30). Reliability of this scale in patients with dementia has been established (31).

The Neuropsychiatric Inventory (NPI), from an interview with the caregiver, was used to assess neuropsychiatric symptoms including delusions, hallucinations, agitation or aggression, depression or dysphoria, anxiety, elation or euphoria, apathy or indifference, disinhibition, irritability or lability, motor disturbance, nighttime behaviors, and appetite and eating changes (23). The Clinical Dementia Rating Scale (CDR) (21) was used to assess clinical stage of dementia.

The Dependence Scale (DS) is a 13-item scale of items found to reflect functional deficiencies important to AD patients and their caregivers (13, 17). It is based on an interview of the caregiver with a trained rater. It is designed to assess assistance that is required of the caregiver rather than the patient’s functional problems. This scale has demonstrated consistency with scales that assess cognitive and functional impairment and robust psychometric properties with strong inter-rater reliability (intraclass correlation of 0.90) and acceptable internal consistency (Cronbach’s alpha of 0.66) in patients with mild-to-moderate AD (13, 17). The Dependence Scale has a higher correlation with the Quality of Life – Alzheimer’s Disease than the CDR or MMSE (32). The DS item “need to be kept company when awake” (yes/no), recording need for supervision while awake, was used as the outcome variable in analysis.

### Data analysis

Descriptive statistics were reported for the sample using number (percent) for categorical variables and median (interquartile range) for continuous variables and stratified by need for supervision while awake. Statistically significant differences between strata were tested using the Wilcoxon rank sum test for continuous variables and the Chi-squared test for categorical variables (Fisher’s exact test was used for categorical variables with > 20% of expected cell counts <5).

We fitted a logistic regression model to describe the association between neuropsychological test results, neuropsychiatric and behavioral symptoms, and EPS with the binary dependent outcome variable recording need for supervision while awake. Missing data were imputed using fully conditional specification. Model results were reported using odds ratios (OR) and 95% confidence intervals (95% CI). All analyses were performed using the *R* statistical software package.

## Results

### Sample characteristics

Table 1 shows characteristics of the sample. Forty-one participants (28.7%) needed supervision while awake, and more than half had depression or dysphoria. Participants were more likely to be Hispanic and female, with a median age of 87 and 6 years of education. In the analyses of the descriptive data (Table 1), patients who needed supervision while awake were more likely to have EPS and hallucinations, and had a lower SRT total Recall score, as compared to those who did not need supervision. Additionally, those who needed supervision were less likely to be Hispanic and more likely to be White, and had fewer years of education, though these differences only approached statistical significance (Table 1).

**Table 1.** Descriptive characteristics for participants, stratified by need for supervision while awake. Abbreviations: EPS = Extrapyramidal signs, IQR = Interquartile range; SRT = Selective Reminding test.

| Characteristic | Overall, N = 143<sup>1</sup> | No, N = 102<sup>1</sup> | Yes, N = 41<sup>1</sup> | p-value |
| --- | --- | --- | --- | --- |
| **Race/Ethnicity** |  |  |  | **0.062** |
| Hispanic | 115 (80%) | 86 (84%) | 29 (71%) |  |
| African-American | 16 (11%) | 11 (11%) | 5 (12%) |  |
| White | 12 (8.4%) | 5 (4.9%) | 7 (17%) |  |
| **Gender** |  |  |  | **0.7** |
| Female | 118 (83%) | 85 (83%) | 33 (80%) |  |
| Male | 25 (17%) | 17 (17%) | 8 (20%) |  |
| **Age (years)** | 87 (83, 91) | 87 (83, 91) | 87 (83, 91) | **0.8** |
| **Education (years)** | 6.0 (3.0, 10.0) | 5.0 (2.0, 10.0) | 8.0 (4.0, 10.0) | **0.057** |
| **Irritability or Lability** | 61 (43%) | 45 (45%) | 16 (39%) | **0.5** |
| Missing | 1 | 1 | 0 |  |
| **Motor Disturbance** | 31 (22%) | 21 (21%) | 10 (24%) | **0.6** |
| Missing | 1 | 1 | 0 |  |
| **Nighttime Behaviors** | 69 (49%) | 47 (47%) | 22 (54%) | **0.4** |
| Missing | 1 | 1 | 0 |  |
| **Appetite and Eating** | 53 (37%) | 36 (36%) | 17 (41%) | **0.5** |
| Missing | 1 | 1 | 0 |  |
| **Delusions** | 57 (40%) | 38 (38%) | 19 (46%) | **0.3** |
| Missing | 1 | 1 | 0 |  |
| **Hallucinations** | 38 (27%) | 22 (22%) | 16 (39%) | **0.035** |
| Missing | 1 | 1 | 0 |  |
| **Agitation or Aggression** | 56 (39%) | 40 (40%) | 16 (39%) | **>0.9** |
| Missing | 1 | 1 | 0 |  |
| **Depression or Dysphoria** | 80 (57%) | 53 (53%) | 27 (66%) | **0.2** |
| Missing | 2 | 2 | 0 |  |
| **Anxiety** | 70 (49%) | 48 (48%) | 22 (54%) | 0.5 |
| Missing | 1 | 1 | 0 |  |
| **Elation or Euphoria** | 13 (9.2%) | 11 (11%) | 2 (4.9%) | 0.3 |
| Missing | 1 | 1 | 0 |  |
| **Apathy or Indifference** | 55 (39%) | 37 (37%) | 18 (44%) | 0.4 |
| Missing | 1 | 1 | 0 |  |
| **Disinhibition** | 39 (27%) | 26 (26%) | 13 (32%) | 0.5 |
| Missing | 1 | 1 | 0 |  |
| **Presence of EPS** | 40 (29%) | 24 (24%) | 16 (41%) | 0.046 |
| Missing | 4 | 2 | 2 |  |
| **SRT Total Recall** | 19.0 (16.0, 23.0) | 20.0 (16.0, 23.0) | 17.0 (13.0, 19.0) | 0.005 |
| Missing | 19 | 9 | 10 |  |
| **Boston Naming Total** | 11.00 (9.00, 12.00) | 11.00 (9.50, 12.00) | 10.50 (8.00, 13.00) | 0.9 |
| Missing | 20 | 11 | 9 |  |
| **CFL Mean** | 4.30 (2.67, 6.15) | 4.00 (2.33, 5.67) | 4.63 (3.07, 7.75) | 0.12 |
| Missing | 32 | 21 | 11 |  |
| **Benton Matching** | 7.00 (5.00, 8.00) | 7.00 (4.00, 8.00) | 6.00 (5.00, 8.00) | >0.9 |
| Missing | 37 | 20 | 17 |  |
| <sup>1</sup>n (%); Median (IQR) |  |  |  |  |
| <sup>2</sup>Fisher's exact test; Pearson's Chi-squared test; Wilcoxon rank sum test |  |  |  |  |

### Logistic regression

The logistic regression model was statistically significant (Likelihood Ratio Test: χ^2^ = 37.1, df = 17, *p* = 0.0032), and showed good to excellent fit (McFadden’s pseudo-r^2^ = 0.22). Table 2 shows logistic model results. CFL mean (OR=1.38, 95% CI [1.15, 1.69]) was positively associated with the need for supervision while awake whereas elation or euphoria (OR=0.12, 95% CI [0.01, 0.68]) and SRT Total Recall (OR=0.88, 95% CI [0.81, 0.96]) were negatively associated with the need for supervision. In other words, those having elation or euphoria had a decreased odds of needing supervision while awake, those with better memory scores had a decreased odds of needing supervision, and those with better letter fluency scores had an increased odds of needing supervision. Additionally, those with hallucinations had an increased odds of needing supervision while awake, though this association only approached statistical significance (OR=2.58, 95% CI [0.90, 7.69]).

**Table 2.** Results from the logistic regression of symptoms associated with the binary dependent outcome variable (need or no need to be kept company when awake on the DS). P-values significant at the 0.05 level are bolded. Abbreviations: CI = Confidence interval; EPS = Extrapyramidal signs; SE = Standard error; SRT = Selective Reminding Test.

| Characteristic | Odds Ratio | SE | 95% CI lower - upper | p-value |
| --- | --- | --- | --- | --- |
| (Intercept) | 1.68 | 1.21 | 0.16, 18.7 | 0.7 |
| Irritability | 0.47 | 0.513 | 0.17, 1.26 | 0.14 |
| Motor disturbance | 0.94 | 0.552 | 0.31, 2.74 | >0.9 |
| Nighttime behaviors | 1.27 | 0.465 | 0.51, 3.18 | 0.6 |
| Appetite and eating | 1.00 | 0.499 | 0.37, 2.66 | >0.9 |
| Delusions | 1.63 | 0.497 | 0.61, 4.35 | 0.3 |
| Hallucinations | 2.58 | 0.544 | 0.90, 7.69 | 0.081 |
| Agitation | 0.59 | 0.517 | 0.21, 1.59 | 0.3 |
| Depression | 1.32 | 0.491 | 0.50, 3.49 | 0.6 |
| Anxiety | 1.03 | 0.509 | 0.37, 2.79 | >0.9 |
| Elation | 0.12 | 0.946 | 0.01, 0.68 | **0.027** |
| Apathy | 1.69 | 0.500 | 0.63, 4.57 | 0.3 |
| Disinhibition | 1.97 | 0.542 | 0.68, 5.76 | 0.2 |
| Presence of EPS | 1.44 | 0.516 | 0.52, 3.97 | 0.5 |
| SRT Total Recall (Immediate) | 0.88 | 0.044 | 0.81, 0.96 | **0.005** |
| Boston Naming Total | 0.89 | 0.087 | 0.75, 1.06 | 0.2 |
| CFL Mean | 1.38 | 0.098 | 1.15, 1.69 | **0.001** |
| Benton Matching | 0.99 | 0.110 | 0.80, 1.23 | >0.9 |

## Discussion

In this analysis of cross-sectional data from a multi-ethnic, community-based cohort study, we found that the need for supervision while awake was significantly associated with the behavioral and neuropsychiatric symptoms of hallucinations and elation. Cognitive measures, including measures of list learning (SRT) and letter fluency (CFL) were also associated with need for supervision while awake. Extrapyramidal symptoms were not significantly associated with need for supervision while awake in the logistic regression.

Our sample for this study is predominantly Hispanic (Table 1). We did not observe a significant effect of race or ethnicity on need for supervision while awake, but in agreement with the previous literature on nursing home placement (13), there was a non-significant trend with the non-Hispanic Whites reporting a higher need for supervision while awake compared to the Hispanic and African-American participants (58% “yes” in White participants compared to 31% in African-Americans and 25% in Hispanics). This could represent a lack of power to detect previously reported associations of race and ethnicity with nursing home placement and/or the differences between need for supervision while awake and nursing home placement. As noted above, nursing home placement as an outcome is likely more influenced than the DS by socioeconomic factors associated with race and ethnicity including availability of unpaid caregivers and resources for paid in-home caregiving and nursing home placement (13).

We found a stronger association between neuropsychiatric and behavioral symptoms and need for supervision while awake, with the highest odds ratio for hallucinations. Patients with active hallucinations often difficulty distinguishing hallucinations from real stimuli and may react to hallucinations in ways that are unsafe, even within minutes of being left alone (e.g., responding violently to hallucinations of people the patient thinks are intruders). While hallucinations and elation are the least common neuropsychiatric and behavioral symptoms of dementia (33), the present study indicates that these symptoms are particularly salient in the important functional outcome of needing supervision while awake and that hallucinations could benefit from targeted non-pharmacological and pharmacological treatment. Unfortunately, antipsychotic medications have the potential for adverse effects of worsened cognition, increased mortality, and extrapyramidal effects (34-36). The effect size of antipsychotic medications for all psychotic symptoms in AD is small, and there is evidence that the efficacy is lower for hallucinations than other psychotic symptoms such as agitation and delusions (34-36). Nonpharmacologic interventions for hallucinations and other psychotic symptoms associated with dementia are indicated (37), and individualized nonpharmacologic treatments for these symptoms are currently under development (38).

Lower list learning performance, associated with poorer memory encoding, was also associated with an increased need for supervision while awake (Table 2). Elation and CFL were also associated with increased need for supervision, but in the other direction. That is, lower elation and higher total CFL score, often associated with frontal lobe function (39), were associated with an increased need for supervision while awake. While this may seem counterintuitive, previous modeling of longitudinal data from the Predictors 1 and 2 studies demonstrated that dependency and mortality are best predicted by inclusion in specific subtypes of patients as defined by changing cognitive, neuropsychiatric, functional, and motor symptoms over time (15). In the current study, only cross-sectional data from the initial visit was analyzed. Thus, initial high elation and low CFL may be associated with a subgroup of patients not initially in need of supervision while awake, but at higher risk for need of supervision while awake later in the course of the illness associated with a subsequent increase in elation and decline in CFL. For example, a patient subgroup with a high prevalence of delusions at initial examination in the Predictors 1 and 2 cohorts (subgroup 3) was at lower risk of need for high-level care three years after initial examination than a patient subgroup with low initial delusions but with subsequent increase in delusions over the next three years (subgroup 2) (15). Future analyses of longitudinal data currently being collected from the Predictors 3 cohort will be important for clarifying the long term impact of neuropsychiatric symptoms and cognitive performance.

In summary, the associations between cognitive, neuropsychiatric, and motor symptoms and need for supervision while awake differ from that of dependency more generally and nursing home placement in instructive ways. These findings provide guidance to clinicians and caregivers regarding predictors of the important symptom of need for supervision while awake, identified by caregivers as a crucial milestone of disease progression, and a strong predictor of increased societal cost of continued care (14). Strengths of the study include a multi-domain longitudinal assessment of an important symptom in an ethnically diverse community-based cohort. A limitation of the study is that it was done cross-sectionally. Further longitudinal research can elucidate predictors of clinically relevant symptoms identified by patients and caregivers with the goal of developing targeted pharmacological and non-pharmacological treatments, a central tenet of patient-centered care.

## Data Availability

All data produced in the present study are available upon reasonable request to the authors.

## Abbreviations

AD: Alzheimer’s disease
ADRD: Alzheimer’s disease and related dementias
MMSE: Mini-Mental State Examination
DS: Dependence Scale (DS)
CDR: Clinical Dementia Rating Scale
SRT: Selective Reminding Test

## Acknowledgements

This study was supported by NIH/NIA (R01AG007370).

